# Radiologically Visible Intraspinal Air Was Very Common After Epidural Access Attempts Among Our Peripartum Patients Who Had Undergone Radiological Examinations

**DOI:** 10.1101/2023.08.15.23294123

**Authors:** Deepak Gupta

**Affiliations:** Wayne State University/Detroit Medical Center, Detroit, Michigan, United States

## Abstract

**Background:** The incidence of intracranial air after neuraxial access attempts was recently quantified. Therefore, the incidence of intraspinal air after neuraxial access attempts needs to be quantified.

**Objective:** To ascertain how common was radiologically visible intraspinal air per computed tomography/magnetic resonance imaging (CT/MR) of spine among our peripartum patients.

**Methods:** For a seven-year period (July 1, 2015-June 30, 2022), medical records of our peripartum patients who had undergone CT/MR of spine were retrospectively reviewed to rule out the presence of recent neuraxial access attempts before their radiological examinations of spine. Concurrently, those radiological examinations of spine were reviewed to rule out the presence of radiologically visible intraspinal air.

**Results:** Among 27 peripartum patients who had undergone CT/MR of spine during the seven-year period, 24 peripartum patients had received recent neuraxial access attempts prior to their radiological examinations wherein 25% had radiologically visible intraspinal air. However, as none of the spinal-only access patients had radiologically visible intraspinal air, intraspinal air was radiologically visible among 32% patients who had recent epidural access attempts.

**Conclusion:** Radiologically visible intraspinal air was very common after epidural access attempts among our peripartum patients who had undergone radiological examinations.

## Introduction

The incidence of radiologically visible intracranial air after neuraxial access attempts was recently quantified among our institution’s peripartum patients spanning a seven-year period [1]. Therefore, the incidence of radiologically visible intraspinal air after neuraxial access attempts needs to be quantified among our institution’s peripartum patients spanning the same seven-year period. It was deemed safe to hypothesize that the retrospective incidence of radiologically visible intraspinal air after neuraxial access attempts would most likely be commoner than the retrospective incidence of radiologically visible intracranial air after neuraxial access attempts considering that neuraxial access attempts especially epidural access attempts involve loss-of-resistance technique which more often than not becomes injection-of-air technique, unknowingly and inadvertently [2-11].

This follow-up completion study was designed to retrospectively ascertain how common was radiologically visible intraspinal air per computed tomography/magnetic resonance imaging (CT/MR) of spine among our institution’s peripartum patients admitted to local Women’s Hospital during the same seven-year period (July 1, 2015-June 30, 2022).

## Methods

After Institutional Review Board approved the follow-up completion study as exempt research, once again the same seven-year period query was re-run by information technology team at our institute but this time to extract the list of peripartum inpatients who had undergone CT/MR of spine. Thereafter, medical records of those patients were reviewed to rule out the presence of recent neuraxial access attempts (epidurals, spinals, combined spinal-epidurals, epidural blood patches) before their radiological examinations of spine. Concurrently, those radiological examinations of spine were reviewed to rule out the presence of radiologically visible intraspinal air.

### Statistical Analysis

2×2-contingency table was once again used to determine relative risk (risk ratio), odds ratio and attributable risk, while online calculator was once again used to determine attributable risk percentage as well as population attributable risk percentage [12-13].

## Results

Over the same seven-year period, only 39 inpatients admitted to peripartum floors had CT/MR of spine but only 27 of those patients were truly peripartum. Among those 27 peripartum patients, 24 peripartum patients had received recent neuraxial access attempts prior to their radiological examinations wherein six peripartum patients (25%) had radiologically visible intraspinal air. As five among those 24 peripartum patients had only received spinals with no documented epidural access attempts and none of these five patients had radiologically visible intraspinal air, the percentage of peripartum patients with radiologically visible intraspinal air increased to 32% among peripartum patients with recent epidural access attempts. Among thirteen peripartum patients with recent epidural access attempts followed by absence of radiologically visible intraspinal air, eleven patients had only received recent epidurals while one patient had received lumbar puncture after epidural and one patient had received epidural blood patch after epidural.

Among the six peripartum patients with radiologically visible intraspinal air, three patients had only received recent epidurals, two patients had received recent epidural blood patches after epidurals and one patient had received recent epidural blood patch after spinal. Interestingly, one patient even went on to receive a second epidural blood patch after the detection of radiologically visible intraspinal air. Among these six peripartum patients with radiologically visible intraspinal air, intraspinal air was radiologically visible in extradural space in four patients, in subdural (intrathecal) space in one patient and in both extradural-subdural (intrathecal) space in one patient. Among these six peripartum patients with radiologically visible intraspinal air, this intraspinal air was radiologically visible in lumbar regions of two patients, in thoracolumbar region of one patient, in cervical region of one patient, and in cervico-thoracic-lumbar regions of two patients.

The statistics data table for risk of intraspinal air with neuraxial access attempts came out as follows:

|  | Number of patients<br>who had<br>radiologically<br>visible intraspinal<br>air | Number of patients<br>who did NOT have<br>radiologically<br>visible intraspinal<br>air | TOTAL |
| --- | --- | --- | --- |
| Number of patients<br>who had neuraxial<br>access attempts<br>before radiological<br>exams | 6 | 18 | 24 |
| Number of patients<br>who did NOT have<br>neuraxial access<br>attempts before<br>radiological exams | 0 | 3 | 3 |
| TOTAL | 6 | 21 | 27 |

Relative risk (risk ratio) for intraspinal air with neuraxial access attempts=(6/24)/(0/3)=Undefined; odds ratio=(6/18)/(0/3)=Undefined; and attributable risk=(6/24)-(0/3)=0.25 with both attributable risk percentage=100% and population attributable risk percentage=100%.

Among those with neuraxial access attempts, the statistics data table for risk of intraspinal air with epidural access attempts came out as follows:

|  | Number of patients<br>who had<br>radiologically<br>visible intraspinal<br>air | Number of patients<br>who did NOT have<br>radiologically<br>visible intraspinal<br>air | TOTAL |
| --- | --- | --- | --- |
| Number of patients<br>who had epidural<br>access attempts<br>before radiological<br>exams | 6 | 13 | 19 |
| Number of patients<br>who did NOT have<br>epidural access<br>attempts before<br>radiological exams | 0 | 5 | 5 |
| TOTAL | 6 | 18 | 24 |

Relative risk (risk ratio) for intraspinal air with epidural access attempts=(6/19)/(0/5)=Undefined; odds ratio=(6/13)/(0/5)=Undefined; and attributable risk=(6/19)-(0/5)=0.32 with both attributable risk percentage=100% and population attributable risk percentage=100%.

## Discussion

Although the number of our institution’s peripartum patients who had undergone CT/MR of spine was very small over the seven-year period, the key finding still was that almost one-third of patients had radiologically visible intraspinal air if they had recent epidural access attempts. The number of our institution’s peripartum patients who had undergone CT/MR of spine being very small deems easy interpretation that our institution’s peripartum patients’ symptoms rarely deemed it necessary to get CT/MR of spine in our institution’s peripartum patients. However, when the radiological examinations had been done, almost one-third of patients had radiologically visible intraspinal air after epidural access attempts.

The study’s limitation was that such small number of peripartum patients receiving CT/MR of spine can preclude the true incidence of radiologically visible intraspinal air after neuraxial access attempts in general and epidural access attempts in particular.

## Conclusion

Radiologically visible intraspinal air was very common after epidural access attempts among our institution’s peripartum patients who had undergone radiological examinations.

## Supporting information

IRB

## Data Availability

All data produced in the present work are contained in the manuscript

## Notes

**Financial Support:** None

**Conflicts of Interests:** None

### Competing Interest Statement

The authors have declared no competing interest.

### Funding Statement

This study did not receive any funding

### Author Declarations

Wayne State University: IRB Administration Office gave ethical approval as concurrence of exemption for this work.

